# Cell type profile variation in menstrual effluent by sample collection method identified using methylation cytometry

**DOI:** 10.1101/2024.03.19.24304465

**Authors:** Irma M. Vlasac, Hannah G. Stolrow, Zaneta M. Thayer, Brock C. Christensen, Luisa Rivera

## Abstract

Menstrual effluent cell profiles have potential as noninvasive biomarkers of female reproductive and gynecological health and disease. We used DNA methylation-based cell type deconvolution (methylation cytometry) to identify cell type profiles in self-collected menstrual effluent. During the second day of their menstrual cycle healthy participants collected menstrual effluent using a vaginal swab, menstrual cup, and pad. Immune cell proportions were highest in menstrual cup samples, and epithelial cells were highest in swab samples. Our work demonstrates the feasibility and utility of menstrual effluent cell profiling in population-level research using remotely collected samples and DNA methylation.

## Introduction

Human females with access to birth control menstruate approximately 400 times during their reproductive lifespan^1^, from menarche in early adolescence around 12 years of age, continuing until menopause at approximately 40-50 years of age. Menstruation is a result of a complex and highly specific system regulated hormonally through the hypothalamus, pituitary, and ovarian endocrine axis, and disruption to the menstrual cycle can be indicative of underlying health issues. As an example, irregular and long menstrual cycles are associated with a range of adverse conditions, including ovarian cancer^2^ and greater risk of premature mortality^3^. Menstrual effluent is comprised of the shed uterine lining developed during the secretory phase of the menstrual cycle as well as cells from the cervix and vagina. As such, menstrual effluent may be a powerful proxy tissue for the endometrium, with recent work finding considerable overlap between menstrual and endometrial cytokine levels, cellular composition, and gene expression profiles^4,5^. Research on menstrual effluent has begun to investigate its potential as an informative biospecimen in women’s health research. Studies to date have focused on elucidating inflammatory profiles and identifying perinatal biomarkers of fertility and endometriosis^5–7^, and as a determinant of adverse pregnancy outcomes. In addition, despite significant disease burden there are no current non-invasive screening tools for ovarian cancer, endometrial cancer, or endometriosis.

Methylation cytometry methods can determine the cell-type admixture of biospecimens with cell reference profiles of DNA methylation and statistical deconvolution. Methylation cytometry methods exist for multiple biospecimen types but have not been applied to menstrual effluent. The hierarchical tumor immune microenvironment epigenetic deconvolution (HiTIMED) algorithm quantifies up to 17 cell types and can be used to estimate cell types in both tumor and non-tumor normal tissues^8^. HiTIMED developed 20 tissue-specific cell reference deconvolution libraries, including endometrial tissue, from which menstrual effluent is shed directly from the endometrium of the uterus. We demonstrate the utility and feasibility of DNA-based cell type profiling in menstrual effluent in healthy females across three collection methods: menstrual cup, menstrual pad, and vaginal swab.

## Materials and Methods

### Study Participants and Samples

Healthy study participants (n=12) were recruited through campus flyers, online communities, and in-person recruitment as part of an IRB-approved study at Dartmouth College. Written consent was obtained, and study participants were screened based on the following eligibility criteria: 18 years of age and older, regular menstrual cycles, no current use of hormonal birth control, no history of reproductive disorders (PCOS, endometriosis, adenomyosis, gynecologic cancers), and no immunomodulatory therapy. Participants completed two questionnaires: health history, and menstrual questionnaire completed at the time of sample collection. Study kits were mailed to participants and contained supplies for menstrual pad, vaginal swab, and menstrual cup self-collection along with instructions to minimize variability among collection methods. Participants were instructed to collect samples on the second day of their period, defined as the second day following the onset of bleeding.

For swab collection, participants were instructed to insert the swab (*Puritan HydraFlock*, #25-3406-H) into their vagina and hold for 15 seconds or until completely saturated. The swab was placed in a 4 mL self-standing centrifuge tube containing 2 mL of DNA/RNA shield (*Zymo*, #R1100-250). This process was repeated twice for the collection of two swabs. To collect the pad sample, participants were provided with an organic cotton liner (natracare, #3145), with a cut 2 × 2-inch square liner secured on top. Participants were instructed to wear the liner until the square was completely saturated, which was estimated to take approximately 5 minutes but may have varied amongst participants.

Once the square was saturated, using the provided tweezers, participants removed the saturated square and placed it in a 4 mL self-standing centrifuge tube containing 2 mL of DNA/RNA shield (Zymo, # R1100-250). The menstrual cup (NeoProMedical, #B092DTN2XS) was worn for sample collection after which a disposable pipette was used to aliquot 2 mL of menstrual effluent into a 4 mL self-standing centrifuge tube containing 2 mL of DNA/RNA shield (Zymo, #R1100-250). Participants were instructed to wear nitrile gloves for all collection modes and place samples in a biohazard collection bag before packaging them in the return box.

### DNA Extraction and Methylation Analysis

DNA was extracted from menstrual cup samples using Qiagen DNeasy Blood and Tissue kit protocol, following manufacturers protocol for extraction from blood samples. DNA was extracted from menstrual pads by cutting a 1x1 centimeter piece of the saturated menstrual pad sample, then using it as the base material for extraction using the Qiagen DNA Investigator’s Kit. DNA was extracted from vaginal swab samples by removing the swab from the plastic swab base, then using the swab material as the base material for extraction using the Qiagen DNA Investigator’s Kit. DNA was extracted from a total of 36 samples, three samples per participant with 12 participants assessed in total. Extracted DNA samples were bisulfite modified using the EZ DNAm Kit (Zymo Research) following the manufacturers optimized protocol for Illumina Methylation arrays^9^. Bisulfite converted DNA was arrayed on the Infinium MethylationEPIC BeadChip V2 array at the Dartmouth Cancer Center Genomics and Molecular Biology Shared Resource.

### Data Processing and Quality Control

MethylationEPICv2 Intensity data (IDATs) were preprocessed for normal exponential out-of-band (Noob) correction using Minfi^10^. Quality control was performed using a detection P, CpG, and sample threshold of 5%, with zero samples demonstrating a percentage of low quality CpG values greater than 5%. Beta value technical replicates were collapsed by averaging probes with common probe ID prefixes using SeSAMe^11^. Single nucleotide polymorphisms (SNPs), probes were used as a quality control metric to assess concordance amongst sample type and participant. Non-CpG cytosine methylation, cancer somatic mutation probes, and probes located on sex chromosomes were filtered. Beta values were normalized using beta mixture quantile normalization (BMIQ). For quality control, sample matching was confirmed using SNP probes.

### Statistical Analysis

Cell proportions were estimated using Hierarchical Immune Tumor Deconvolution (HiTIMED)^8^, designated at deconvolution level four, set to non-tumor normal tissue with the uterine carcinoma deconvolution library. Cell proportions from each sample type (menstrual pad, menstrual cup, vaginal swab) were assessed for significant differences between the three collection sample types using a Kruskal-Wallis test. Unsupervised hierarchical clustering was performed using the top 20,000 most variable CpGs. A Recursively Partitioned Mixture Model (RPMM)^12^ was fit using the 20,000 most variably methylated CpGs to identify subgroups within the methylation data. Association of RPMM cluster membership with sample collection method was tested with a Fisher’s exact test.

## Results

DNA methylation data from each collection method matched to a total of twelve study participants (**Supplementary Table 1**) was visualized with unsupervised hierarchical clustering of the 20,000 most variably methylated CpGs (**Figure 1A**). To test associations of methylation with sample collection method, a recursively partitioned mixture model was fit to the 20,000 most variably methylated CpGs (**Figure 1B**), resulting in four clusters. The majority of cup samples (75%) were in cluster rRR, and sample collection method was significantly associated with methylation cluster membership (Fisher’s Exact P=3.9E-05, **Figure 1C**).

**Figure 1.**
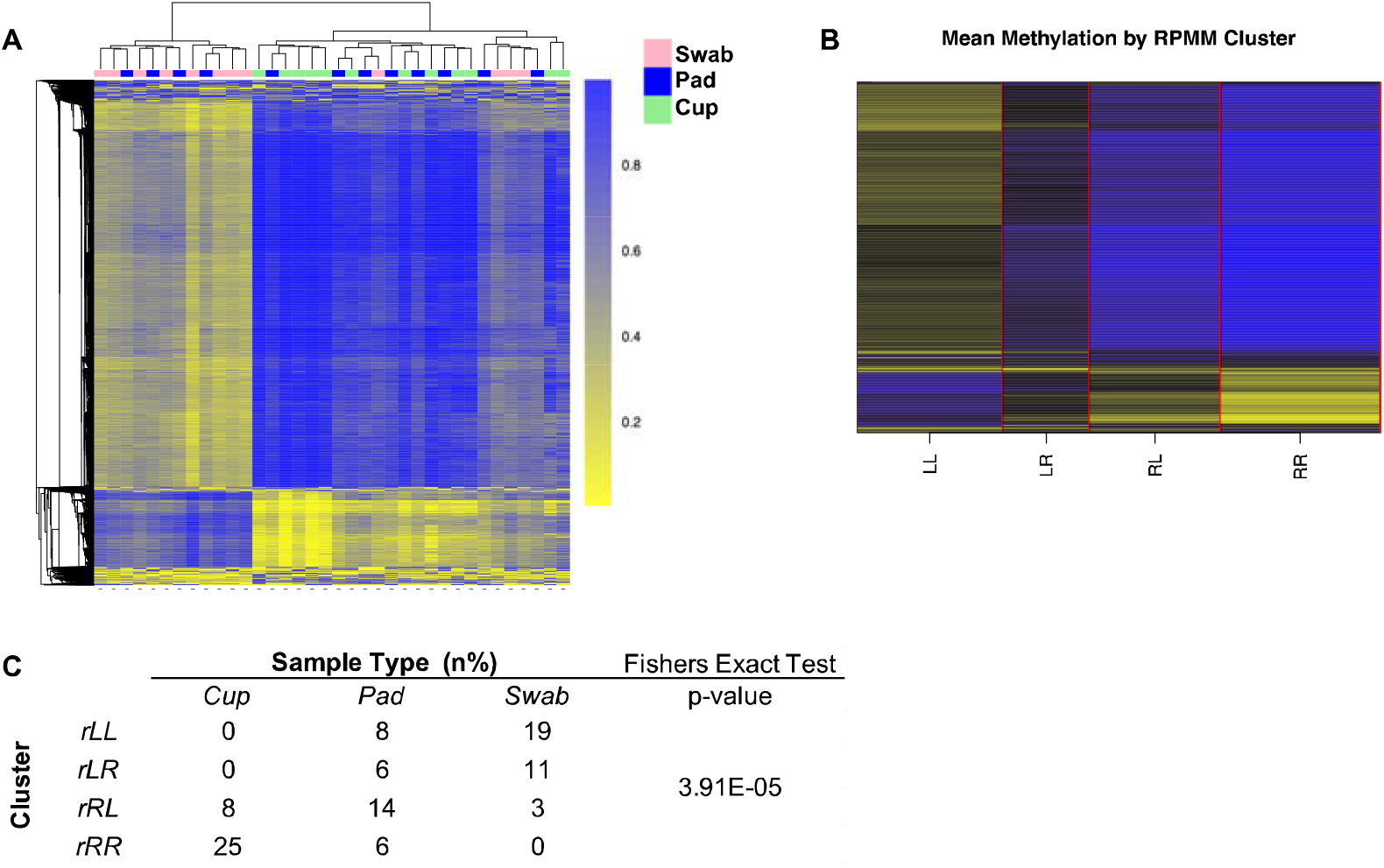
A) Unsupervised hierarchical clustering based on top 20,000 most variable CpGs. B) Recursively Partitioned Mixture Model (RPMM) with mean methylation displayed in each of the four clusters. C) table of RPMM cluster membership by sample type.

Menstrual effluent cell type proportions were estimated using HiTIMED in all samples and compared among collection methods. Epithelial and stromal cell proportions were significantly higher in swab samples compared to pad samples, with cup samples demonstrating the lowest epithelial and stromal cell proportions; cup samples had significantly higher granulocyte, natural killer, and T cell proportions compared to swab samples (P <0.05, Kruskal-Wallis test, **Figure 2**).

**Figure 2.**
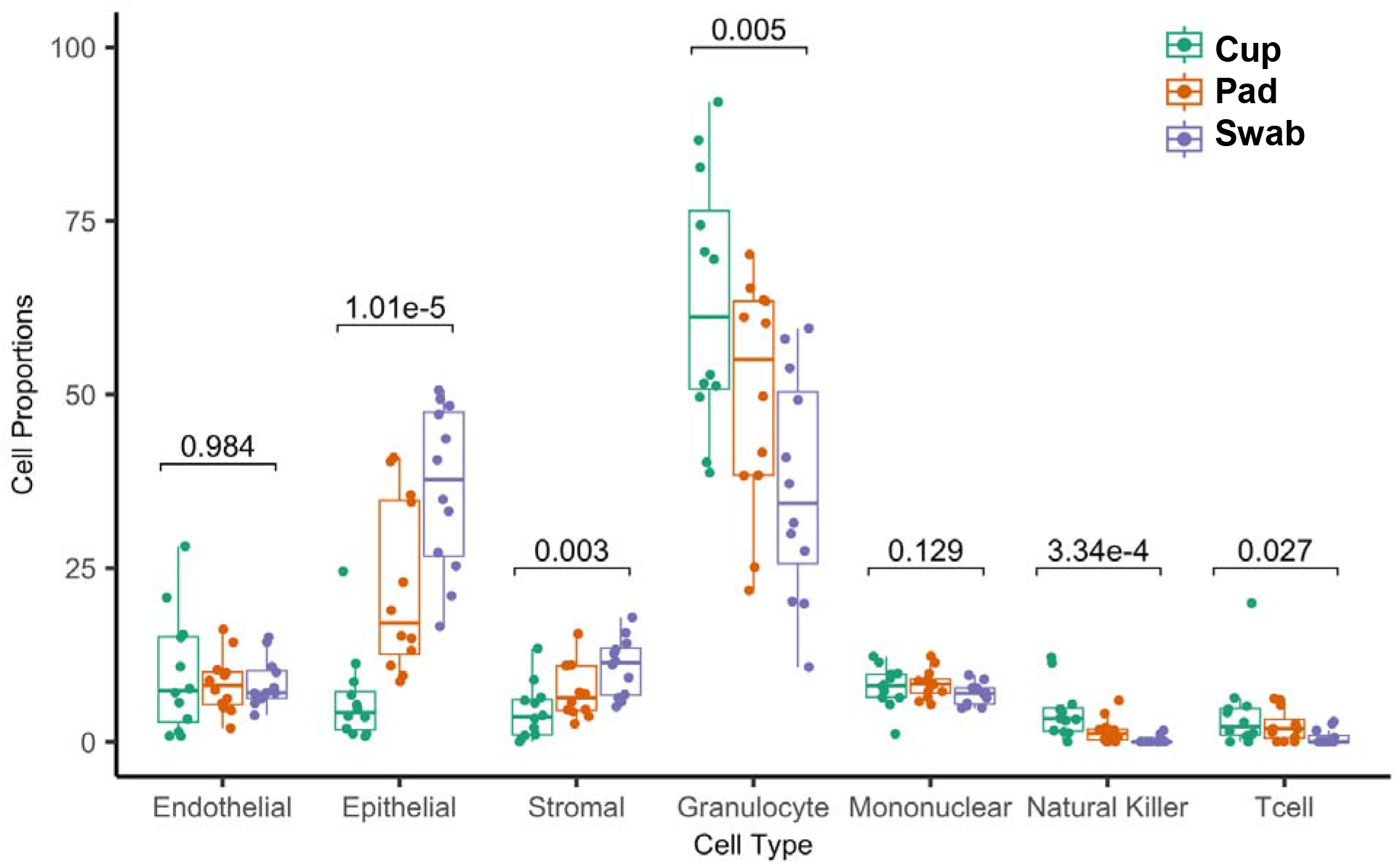
Menstrual effluent cell type proportions differ among menstrual cup, menstrual pad, and vaginal swab collection methods as determined by methylation cytometry.

## Discussion

Our work demonstrates the utility of cell type profiling in menstrual effluent with methylation cytometry for research applications in female reproductive and gynecological health and disease. Methylation cytometry cell deconvolution analysis identified cell type proportion differences by sample collection method. Menstruation is a process of tissue inflammation and is marked by an increase in myeloid immune cells and leukocytes recruited in response to inflammation from necrotizing and shedding tissue^13,14^. Menstrual cup samples had the highest immune cell proportions, specifically granulocyte, natural killer, and T cell proportions in comparison to menstrual pad and vaginal swab samples. Considering diseases marked by abnormally high inflammation, such as endometriosis, future studies utilizing menstrual effluent should consider collection method as it relates with the study goals and hypotheses. Studies aimed at elucidating immune cell profiles with gynecologic health and disease may be better powered to discern effects with menstrual cup and pad collection methods. Similarly, studies which may be more focused on assessing conditions that affect epithelial and stromal cells of the cervix or vagina, may favor menstrual pads or vaginal swabs for sample collection. In any case, application of methylation cytometry methods enables investigation of cell-type-dependent and cell-type-independent associations with health and disease.

We expected cell type proportion differences based on collection method similarly to how we observed them. The vaginal swab collection method had higher epithelial cell proportions, which is likely related to the method of sample collection considering the participant collected the sample shallowly within the vaginal canal as opposed to closer to the cervix. Similarly, the menstrual pad collection likely demonstrates higher epithelial cell proportions considering the shed menstrual effluent travels through the vaginal canal and vulva before being absorbed on the menstrual pad. While menstrual cup sample collections demonstrate the highest immune cell proportions compared to other sample collection methods, menstrual effluent collected by menstrual pad holds great promise for future research studies given it captures cell types from the immune, epithelial, and stromal cell compartments. Considering menstrual pad collection is the only method of collection that doesn’t require insertion, it allows for a truly non-invasive, non-penetrative sample collection, allowing for sample collection from adolescents and individuals that may not want to use an insertional collection method. As menstrual pads can easily and inexpensively be shipped in the mail and are widely available, they may provide greater potential in the scope of population level studies and studies conducted in resource limited populations. Use of DNA-based cell typing in menstrual effluent samples combined with remote, noninvasive sampling methods potentiate both research and future clinical applications of menstrual effluent specimens that benefit women’s health.

## Data Availability

All data produced in the present study are available upon reasonable request to the authors.

---

**Supplementary Table 1.**

| Characteristic | Participants (N = 12) |
| --- | --- |
| **Age (%)** |  |
| 18 - 25 | 7 (58.3) |
| 26 - 35 | 2 (16.7) |
| > 35 | 3 (25.0) |
| *Average Age* | 27 |
| **Parity (%)** |  |
| Nulliparous | 8 (66.7) |
| Multiparous | 4 (33.3) |
| **Menstrual Pain (%)** |  |
| Not painful | 1 (8.3) |
| A little painful | 4 (33.3) |
| Somewhat painful | 6 (50.0) |
| Very painful | 1 (8.3) |
| **Recent Illness (%)** |  |
| No | 11 (91.7) |
| Yes | 1 (8.3) |
| **Pain Medication (%)** |  |
| None | 9 (75.0) |
| Ibuprofen | 3 (25.0) |

## Declarations

### Ethics approval and consent to participate

### Availability of data and materials

### Author’s contributions

LR and ZMT designed the study. IMV and HGS processed samples. IMV performed the analysis and drafted the manuscript. All authors contributed to and approved the final manuscript. Thank you to Claire Pingitore and Sovi Mya-Wellons for their assistance in assembling and sending study kits.

